## Supplementary information for "Socio-demographic and genetic risk factors for drug adherence and persistence: a retrospective nationwide and biobank study across 5 medication classes and 1 845 665 individuals"

### Supplementary Note

#### FinnGen

*Genotyping and quality control*

FinnGen consists of prospectively recruited samples and a series of legacy cohorts with genotypes already available. Prospective samples were genotyped using the ThermoFisher Axiom custom array which tags a total of 655,973 variants. Genotype calling was performed using the Array Power Tools software. Legacy cohorts were genotyped using various Illumina arrays and genotype calling was performed using either GenCall or zCall algorithms. For both prospective and legacy cohorts the following quality control metrics were used.

Samples were removed if:

- Pihat was > 0.9 and the samples were not monozygotic or replicates
- There was a discrepancy between reported sex and genetically determined sex (F-value ≤ 0.3 for females and ≥ 0.8 for males)
- Missingness was ≥ 5%
- Heterozygosity was ±4 standard deviations from the population average
- Pihat was > 0.1 with 14 or more samples
- Samples were ±4 standard deviations away from the population average according to the first two genetic principal components.
- Samples were tagged should there be evidence of a mendelian error or contain replicate samples with over 50,000 discrepancies.

Variants were removed if:

- The variant failed the Hardy-Weinberg Equilibrium test (p-value < 10-6)
- The variant had a call rate < 98%

*Imputation*

Pre-phasing was performed using Eagle 2.3.5 and samples were imputed using the SiSu v3 imputation reference panel. This reference panel is specific to the Finnish population, containing high-coverage (25-30x) whole-genome sequencing data from 3,775 Finns and 16,962,023 variants with minor allele count ≥ 3. After imputation, 16,387,711 variants were imputed with high quality (INFO > 0.6).

*Ancestry assignment*

Firstly, the FinnGen samples were combined with the 1000 genomes phase 3 dataset. Genetic principal components were calculated using a subset of 49,451 pruned SNPs. Aberrant was used to identify and remove samples that deviated from the main cluster. A probability of belonging to either a North-Western European or Finnish population was calculated by firstly performing PCA with individuals belonging to these ancestries from 1000 genomes data. FinnGen samples were then projected onto this PCA space and Mahalanobis distances calculated for each sample against each of the two ancestries. Samples were retained if there was ≥ 95% probability of belonging to the Finnish ancestry cluster.

**Estonian Biobank**

*Genotyping and quality control*

Genotyping of DNA samples from the Estonian Biobank was done at the Core Genotyping Lab of the Institute of Genomics, University of Tartu using the Illumina Global Screening Arrays (GSAv1.0, GSAv2.0, and GSAv2.0_EST). Altogether 206,448 samples were genotyped and then PLINK format files were created using Illumina GenomeStudio v2.0.4. During the quality control all individuals with call-rate < 95% or mismatching sex that was defined based on the heterozygosity of X chromosome and sex in the phenotype data, were excluded from the analysis. Variants were filtered by call-rate < 95% and HWE p-value < 1e-4 (autosomal variants only). Variant positions were updated to Genome Reference Consortium Human Build 37 and all variants were changed to be from TOP strand using reference information provided by Dr. Will Rayner from the University of Oxford (https://www.well.ox.ac.uk/~wrayner/strand/). After QC the dataset contained 202,910 samples for imputation.

*Imputation*

Before imputation variants with MAF<1% and Indels were removed. Prephasing was done using the Eagle v2.3 software [1] (number of conditioning haplotypes Eagle2 uses when phasing each sample was set to: --Kpbwt=20000) and imputation was carried out using Beagle v.18May20.d20 [2,3] with an effective population size ne=20,000. As a reference, Estonian population specific imputation reference of 2297 WGS samples was used [4].

*Ancestry assignment*

Further, EstBB samples were combined with the 1000 genomes phase 3 dataset for ancestry analysis. Genetic principal components were calculated using a subset of quality controlled and pruned genotyped SNPs. This was further used to identify and remove samples that deviated from the main cluster.

### Effect of polytherapy on adherence and persistence

We assessed the effect of polytherapy on drug adherence and persistence with respect to the five medications defined in the primary analysis. For each medication, we determined if any of the other four treatments were concurrent in the following manner: for adherence, we considered a medication regimen concurrent if the time between the first and last purchase recorded was overlapping at any time with the timespan used for adherence calculation; for persistence if the time between first and last purchase recorded of the potential concurrent treatment contained the purchase date used for persistence calculation. We fitted a linear model for persistence and adherence with a categorical variable with three levels (0 for no concurrent treatment, 1 for one concurrent treatment, and 2 for more than one concurrent treatment), adjusting for the baseline covariates used in the primary analysis (**Health and socio-demographic risk factors for persistence and adherence**). The percentages of change in adherence for polytherapy are reported and OR for persistence are reported in Supplementary Tables 21-22.
We observed that the presence of at least once concurrent treatment was consistently associated with both increased adherence and higher odds of persistence. The percentage increase in adherence with one concurrent treatment ranged from 0.6% (blood pressure medications) to 3.6% (antiplatelets). The ORs of being persistent between one concurrent treatment and no concurrent treatments go from 1.04 (not statistically significant for breast cancer medications) to 2.79 (anticoagulants). Moreover, we find consistently larger effect sizes for two or more concurrent treatments compared with only one concurrent treatment, except for blood pressure medications.

### Characterization of variants genome-wide significantly associated with adherence and persistence

Supplementary Table 12 reports the 4 variants associated with either adherence or persistence at $P < 5 \times 10^{-8}$. We further characterized of these variants based on evidences reported in Open Target Genetics [5].

- *rs1339882991,* positively associated with both adherence and persistence to BP medications, is an intronic variant located in proximity of the *WNT2B* gene, showing a V2G assignment to the same gene based on evidence from brain tissue eQTLs. This variant was previously reported to be associated with increased risk of hypertension[6] and higher blood pressure[7].
- *rs111349244,* associated with lower odds of persistence to BP medications, is an intronic variant located near the *LINC02227* gene and was associated with a lower number of antihypertensive medication purchases and decreased risk of hypertension in FinnGen.
- *rs12149025,* associated with lower persistence to DOAC, is an upstream gene variant located in proximity of the *CBFA2T3* gene, with V2G assignment to *CDH15* based on PCHi-C[8] evidences and evidences from muscle tissue eQTLs.

- *rs548379361,* associated with lower adherence to breast cancer medications, is an intronic variant near the *CFAP44* gene.

### Supplementary Figures

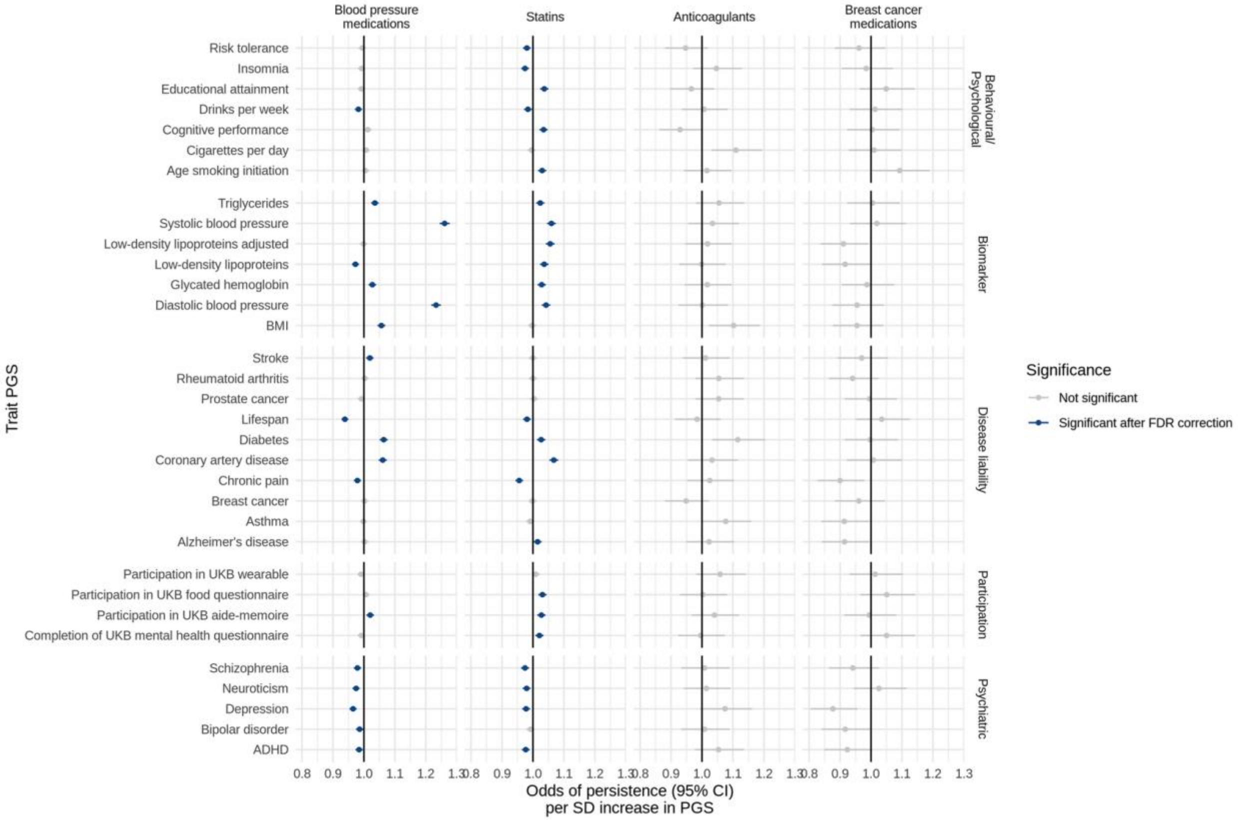


**Supplementary Figure 1 | Associations between persistence (as defined for the sensitivity analysis) and PGS for 33 clinically relevant traits.** Odds of persistence per 1-SD increase in trait PGS. ORs are from a logistic regression model adjusted for sex, age at initiation, first 10 genetic principal components. Error bars represent the 95% confidence interval for the estimates. Results are reported in Supplementary Table 18.

### FinnGen Contributors

### Steering Committee


| | |
|---|---|
| Aarno Palotie | Institute for Molecular Medicine Finland (FIMM), HiLIFE, University of Helsinki, Helsinki, Finland; B Institute of MIT and Harvard; Massachusetts General Hospital |
| Mark Daly | Institute for Molecular Medicine Finland (FIMM), HiLIFE, University of Helsinki, Helsinki, Finland; Broad Institute of MIT and Harvard; Massachusetts General Hospital |

**Pharmaceutical Companies**

| | |
|---|---|
| Bridget Riley-Gills | Abbvie, Chicago, IL, United States |
| Howard Jacob | Abbvie, Chicago, IL, United States |
| Dirk Paul | Astra Zeneca, Cambridge, United Kingdom |
| Athena Matakidou | Astra Zeneca, Cambridge, United Kingdom |
| Adam Platt | Astra Zeneca, Cambridge, United Kingdom |
| Heiko Runz | Biogen, Cambridge, MA, United States |
| Sally John | Biogen, Cambridge, MA, United States |
| George Okafo | Boehringer Ingelheim, Ingelheim am Rhein, Germany |
| Nathan Lawless | Boehringer Ingelheim, Ingelheim am Rhein, Germany |
| Robert Plenge | Bristol Myers Squibb, New York, NY, United States |
| Joseph Maranville | Bristol Myers Squibb, New York, NY, United States |
| Mark McCarthy | Genentech, San Francisco, CA, United States |
| Julie Hunkapiller | Genentech, San Francisco, CA, United States |
| Margaret G. Ehm | GlaxoSmithKline, Collegeville, PA, United States |
| Kirsi Auro | GlaxoSmithKline, Espoo, Finland |
| Simonne Longerich | Merck, Kenilworth, NJ, United States |
| Caroline Fox | Merck, Kenilworth, NJ, United States |
| Anders Mälarstig | Pfizer, New York, NY, United States |
| Katherine Klinger | Translational Sciences, Sanofi R&D, Framingham, MA, USA |
| Deepak Raipal | Translational Sciences, Sanofi R&D, Framingham, MA, USA |
| Eric Green | Maze Therapeutics, San Francisco, CA, United States |
| Robert Graham | Maze Therapeutics, San Francisco, CA, United States |

| | |
|---|---|
| Robert Yang | Janssen Biotech, Beerse, Belgium |
| Chris O´Donnell | Novartis Institutes for BioMedical Research, Cambridge, MA, United States |

**University of Helsinki & Biobanks**

| | |
|---|---|
| Tomi Mäkelä | HiLIFE, University of Helsinki, Finland, Finland |
| Jaakko Kaprio | Institute for Molecular Medicine Finland (FIMM), HiLIFE, University of Helsinki, Helsinki, Finland |
| Petri Virolainen | Auria Biobank / University of Turku / Hospital District of Southwest Finland, Turku, Finland |
| Antti Hakanen | Auria Biobank / University of Turku / Hospital District of Southwest Finland, Turku, Finland |
| Terhi Kilpi | THL Biobank / Finnish Institute for Health and Welfare (THL), Helsinki, Finland |
| Markus Perola | THL Biobank / Finnish Institute for Health and Welfare (THL), Helsinki, Finland |
| Jukka Partanen | Finnish Red Cross Blood Service / Finnish Hematology Registry and Clinical Biobank, Helsinki, Finland |
| Anne Pitkäranta | Helsinki Biobank / Helsinki University and Hospital District of Helsinki and Uusimaa, Helsinki |
| Juhani Junttila | Northern Finland Biobank Borealis / University of Oulu / Northern Ostrobothnia Hospital District, Oulu, Finland |
| Raisa Serpi | Northern Finland Biobank Borealis / University of Oulu / Northern Ostrobothnia Hospital District, Oulu, Finland |
| Tarja Laitinen | Finnish Clinical Biobank Tampere / University of Tampere / Pirkanmaa Hospital District, Tampere, Finland |
| Veli-Matti Kosma | Biobank of Eastern Finland / University of Eastern Finland / Northern Savo Hospital District, Kuopio, Finland |
| Jari Laukkanen | Central Finland Biobank / University of Jyväskylä / Central Finland Health Care District, Jyväskylä, Finland |
| Marco Hautalahti | FINBB - Finnish biobank cooperative |

**Other Experts/ Non-Voting Members**

| | |
|---|---|
| Outi Tuovila | Business Finland, Helsinki, Finland |
| Raimo Pakkanen | Business Finland, Helsinki, Finland |

### Scientific Committee

**Pharmaceutical companies**

| | |
|---|---|
| Jeffrey Waring | Abbvie, Chicago, IL, United States |
| Bridget Riley-Gillis | Abbvie, Chicago, IL, United States |
| Fedik Rahimov | Abbvie, Chicago, IL, United States |
| Ioanna Tachmazidou | Astra Zeneca, Cambridge, United Kingdom |
| Chia-Yen Chen | Biogen, Cambridge, MA, United States |

| | |
|---|---|
| Heiko Runz | Biogen, Cambridge, MA, United States |
| Zhihao Ding | Boehringer Ingelheim, Ingelheim am Rhein, Germany |
| Marc Jung | Boehringer Ingelheim, Ingelheim am Rhein, Germany |
| Shameek Biswas | Bristol Myers Squibb, New York, NY, United States |
| Rion Pendergrass | Genentech, San Francisco, CA, United States |

154

| | |
|---|---|
| Julie Hunkapiller | Genentech, San Francisco, CA, United States |
| Margaret G. Ehm | GlaxoSmithKline, Collegeville, PA, United States |
| David Pulford | GlaxoSmithKline, Stevenage, United Kingdom |
| Neha Raghavan | Merck, Kenilworth, NJ, United States |
| Adriana Huertas-Vazquez | Merck, Kenilworth, NJ, United States |
| Jae-Hoon Sul | Merck, Kenilworth, NJ, United States |
| Anders Mälarstig | Pfizer, New York, NY, United States |
| Xinli Hu | Pfizer, New York, NY, United States |
| Katherine Klinger | Translational Sciences, Sanofi R&D, Framingham, MA, USA |
| Robert Graham | Maze Therapeutics, San Francisco, CA, United States |
| Eric Green | Maze Therapeutics, San Francisco, CA, United States |
| Sahar Mozaffari | Maze Therapeutics, San Francisco, CA, United States |
| Dawn Waterworth | Janssen Research & Development, LLC, Spring House, PA, United States |
| Nicole Renaud | Novartis Institutes for BioMedical Research, Cambridge, MA, United States |
| Ma´en Obeidat | Novartis Institutes for BioMedical Research, Cambridge, MA, United States |

**University of Helsinki & Biobanks**


| | |
|---|---|
| Samuli Ripatti | Institute for Molecular Medicine Finland (FIMM), HiLIFE, University of Helsinki, Helsinki, Finland |
| Johanna Schleutker | Auria Biobank / Univ. of Turku / Hospital District of Southwest Finland, Turku, Finland |
| Markus Perola | THL Biobank / Finnish Institute for Health and Welfare (THL), Helsinki, Finland |
| Mikko Arvas | Finnish Red Cross Blood Service / Finnish Hematology Registry and Clinical Biobank, Helsinki, Finland |
| Olli Carpén | Helsinki Biobank / Helsinki University and Hospital District of Helsinki and Uusimaa, Helsinki |
| Reetta Hinttala | Northern Finland Biobank Borealis / University of Oulu / Northern Ostrobothnia Hospital District, Oulu, Finland |
| Johannes Kettunen | Northern Finland Biobank Borealis / University of Oulu / Northern Ostrobothnia Hospital District, Oulu, Finland |
| Arto Mannermaa | Biobank of Eastern Finland / University of Eastern Finland / Northern Savo Hospital District, Kuopio, Finland |
| Katriina Aalto-Setälä | Faculty of Medicine and Health Technology, Tampere University, Tampere, Finland |
| Mika Kähönen | Finnish Clinical Biobank Tampere / University of Tampere / Pirkanmaa Hospital District, Tampere, Finland |
| Jari Laukkanen | Central Finland Biobank / University of Jyväskylä / Central Finland Health Care District, Jyväskylä, Finland |
| Johanna Mäkelä | FINBB - Finnish biobank cooperative |


### Clinical Groups

#### Neurology Group


| | |
|---|---|
| Reetta Kälviäinen | Northern Savo Hospital District, Kuopio, Finland |
| Valtteri Julkunen | Northern Savo Hospital District, Kuopio, Finland |
| Hilkka Soininen | Northern Savo Hospital District, Kuopio, Finland |
| Anne Remes | Northern Ostrobothnia Hospital District, Oulu, Finland |
| Mikko Hiltunen | University of Eastern Finland, Kuopio, Finland |
| Jukka Peltola | Pirkanmaa Hospital District, Tampere, Finland |
| Minna Raivio | Hospital District of Helsinki and Uusimaa, Helsinki, Finland |
| Pentti Tienari | Hospital District of Helsinki and Uusimaa, Helsinki, Finland |
| Juha Rinne | Hospital District of Southwest Finland, Turku, Finland |
| Roosa Kallionpää | Hospital District of Southwest Finland, Turku, Finland |

| | |
|---|---|
| Juulia Partanen | Institute for Molecular Medicine Finland, HiLIFE, University of Helsinki, Finland |
| Ali Abbasi | Abbvie, Chicago, IL, United States |
| Adam Ziemann | Abbvie, Chicago, IL, United States |
| Nizar Smaoui | Abbvie, Chicago, IL, United States |
| Anne Lehtonen | Abbvie, Chicago, IL, United States |
| Susan Eaton | Biogen, Cambridge, MA, United States |
| Heiko Runz | Biogen, Cambridge, MA, United States |
| Sanni Lahdenperä | Biogen, Cambridge, MA, United States |
| Shameek Biswas | Bristol Myers Squibb, New York, NY, United States |
| Julie Hunkapiller | Genentech, San Francisco, CA, United States |
| Natalie Bowers | Genentech, San Francisco, CA, United States |
| Edmond Teng | Genentech, San Francisco, CA, United States |
| Rion Pendergrass | Genentech, San Francisco, CA, United States |
| Fanli Xu | GlaxoSmithKline, Brentford, United Kingdom |
| David Pulford | GlaxoSmithKline, Stevenage, United Kingdom |
| Kirsi Auro | GlaxoSmithKline, Espoo, Finland |
| Laura Addis | GlaxoSmithKline, Brentford, United Kingdom |
| John Eicher | GlaxoSmithKline, Brentford, United Kingdom |
| Qingqin S Li | Janssen Research & Development, LLC, Titusville, NJ 08560, United States |
| Karen He | Janssen Research & Development, LLC, Spring House, PA, United States |
| Ekaterina Khramtsova | Janssen Research & Development, LLC, Spring House, PA, United States |
| Neha Raghavan | Merck, Kenilworth, NJ, United States |

**Gastroenterology Group**

| | |
|---|---|
| Martti Färkkilä | Hospital District of Helsinki and Uusimaa, Helsinki, Finland |
| Jukka Koskela | Hospital District of Helsinki and Uusimaa, Helsinki, Finland |
| Sampsa Pikkarainen | Hospital District of Helsinki and Uusimaa, Helsinki, Finland |
| Airi Jussila | Pirkanmaa Hospital District, Tampere, Finland |
| Katri Kaukinen | Pirkanmaa Hospital District, Tampere, Finland |
| Timo Blomster | Northern Ostrobothnia Hospital District, Oulu, Finland |

| | |
|---|---|
| Mikko Kiviniemi | Northern Savo Hospital District, Kuopio, Finland |
| Markku Voutilainen | Hospital District of Southwest Finland, Turku, Finland |
| Mark Daly | Institute for Molecular Medicine, Finland (FIMM), HiLIFE, University of Helsinki, Helsinki, Finland; Broad Institute of MIT and Harvard; Massachusetts General Hospital |
| Ali Abbasi | Abbvie, Chicago, IL, United States |
| Jeffrey Waring | Abbvie, Chicago, IL, United States |
| Nizar Smaoui | Abbvie, Chicago, IL, United States |
| Fedik Rahimov | Abbvie, Chicago, IL, United States |
| Anne Lehtonen | Abbvie, Chicago, IL, United States |
| Tim Lu | Genentech, San Francisco, CA, United States |
| Natalie Bowers | Genentech, San Francisco, CA, United States |
| Rion Pendergrass | Genentech, San Francisco, CA, United States |
| Linda McCarthy | GlaxoSmithKline, Brentford, United Kingdom |
| Amy Hart | Janssen Research & Development, LLC, Spring House, PA, United States |
| Meijian Guan | Janssen Research & Development, LLC, Spring House, PA, United States |
| Jason Miller | Merck, Kenilworth, NJ, United States |
| Kirsi Kalpala | Pfizer, New York, NY, United States |
| Melissa Miller | Pfizer, New York, NY, United States |
| Xinli Hu | Pfizer, New York, NY, United States |


**Rheumatology Group**


| | |
|---|---|
| Kari Eklund | Hospital District of Helsinki and Uusimaa, Helsinki, Finland |
| Antti Palomäki | Hospital District of Southwest Finland, Turku, Finland |
| Pia Isomäki | Pirkanmaa Hospital District, Tampere, Finland |
| Laura Pirilä | Hospital District of Southwest Finland, Turku, Finland |
| Oili Kaipiainen-Seppänen | Northern Savo Hospital District, Kuopio, Finland |
| Johanna Huhtakangas | Northern Ostrobothnia Hospital District, Oulu, Finland |
| Nina Mars | Institute for Molecular Medicine Finland (FIMM), HiLIFE, University of Helsinki, Helsinki, Finland |
| Ali Abbasi | Abbvie, Chicago, IL, United States |
| Jeffrey Waring | Abbvie, Chicago, IL, United States |
| Fedik Rahimov | Abbvie, Chicago, IL, United States |

| | |
|---|---|
| Apinya Lertratanakul | Abbvie, Chicago, IL, United States |
| Nizar Smaoui | Abbvie, Chicago, IL, United States |
| Anne Lehtonen | Abbvie, Chicago, IL, United States |
| David Close | Astra Zeneca, Cambridge, United Kingdom |
| Marla Hochfeld | Bristol Myers Squibb, New York, NY, United States |
| Natalie Bowers | Genentech, San Francisco, CA, United States |
| Rion Pendergrass | Genentech, San Francisco, CA, United States |
| Jorge Esparza Gordillo | GlaxoSmithKline, Brentford, United Kingdom |
| Kirsi Auro | GlaxoSmithKline, Espoo, Finland |
| Dawn Waterworth | Janssen Research & Development, LLC, Spring House, PA, United States |
| Fabiana Farias | Merck, Kenilworth, NJ, United States |
| Kirsi Kalpala | Pfizer, New York, NY, United States |
| Nan Bing | Pfizer, New York, NY, United States |
| Xinli Hu | Pfizer, New York, NY, United States |

**Pulmonology Group**

| | |
|---|---|
| Tarja Laitinen | Pirkanmaa Hospital District, Tampere, Finland |
| Margit Pelkonen | Northern Savo Hospital District, Kuopio, Finland |
| Paula Kauppi | Hospital District of Helsinki and Uusimaa, Helsinki, Finland |
| Hannu Kankaanranta | University of Gothenburg, Gothenburg, Sweden/ Seinäjoki Central Hospital, Seinäjoki, Finland/ Tampere University, Tampere, Finland |
| Terttu Harju | Northern Ostrobothnia Hospital District, Oulu, Finland |
| Riitta Lahesmaa | Hospital District of Southwest Finland, Turku, Finland |
| Nizar Smaoui | Abbvie, Chicago, IL, United States |
| Alex Mackay | Astra Zeneca, Cambridge, United Kingdom |
| Glenda Lassi | Astra Zeneca, Cambridge, United Kingdom |
| Susan Eaton | Biogen, Cambridge, MA, United States |
| Hubert Chen | Genentech, San Francisco, CA, United States |
| Rion Pendergrass | Genentech, San Francisco, CA, United States |
| Natalie Bowers | Genentech, San Francisco, CA, United States |
| Joanna Betts | GlaxoSmithKline, Brentford, United Kingdom |

Kirsi Auro GlaxoSmithKline, Espoo, Finland
Rajashree Mishra GlaxoSmithKline, Brentford, United Kingdom
Majd Mouded Novartis, Basel, Switzerland
Debby Ngo Novartis, Basel, Switzerland

**Cardiometabolic Diseases Group**

Teemu Niiranen Finnish Institute for Health and Welfare (THL), Helsinki, Finland
Felix Vaura Finnish Institute for Health and Welfare (THL), Helsinki, Finland
Veikko Salomaa Finnish Institute for Health and Welfare (THL), Helsinki, Finland
Kaj Metsärinne Hospital District of Southwest Finland, Turku, Finland
Jenni Aittokallio Hospital District of Southwest Finland, Turku, Finland
Mika Kähönen Pirkanmaa Hospital District, Tampere, Finland
Jussi Hernesniemi Pirkanmaa Hospital District, Tampere, Finland
Daniel Gordin Hospital District of Helsinki and Uusimaa, Helsinki, Finland
Juha Sinisalo Hospital District of Helsinki and Uusimaa, Helsinki, Finland
Marja-Riitta Taskinen Hospital District of Helsinki and Uusimaa, Helsinki, Finland
Tiinamaija Tuomi Hospital District of Helsinki and Uusimaa, Helsinki, Finland
Timo Hiltunen Hospital District of Helsinki and Uusimaa, Helsinki, Finland
Jari Laukkanen Central Finland Health Care District, Jyväskylä, Finland
Amanda Elliott Institute for Molecular Medicine Finland (FIMM), HiLIFE, University of Helsinki, Helsinki, Finland; Broad Institute, Cambridge, MA, USA and Massachusetts General Hospital, Boston, MA, USA
Mary Pat Reeve Institute for Molecular Medicine Finland (FIMM), HiLIFE, University of Helsinki, Helsinki, Finland
Sanni Ruotsalainen Institute for Molecular Medicine Finland (FIMM), HiLIFE, University of Helsinki, Helsinki, Finland
Benjamin Challis Astra Zeneca, Cambridge, United Kingdom
Dirk Paul Astra Zeneca, Cambridge, United Kingdom
Julie Hunkapiller Genentech, San Francisco, CA, United States
Natalie Bowers Genentech, San Francisco, CA, United States
Rion Pendergrass Genentech, San Francisco, CA, United States
Audrey Chu GlaxoSmithKline, Brentford, United Kingdom
Kirsi Auro GlaxoSmithKline, Espoo, Finland
Dermot Reilly Janssen Research & Development, LLC, Boston, MA, United States

| | |
|---|---|
| Mike Mendelson | Novartis, Boston, MA, United States |
| Jaakko Parkkinen | Pfizer, New York, NY, United States |
| Melissa Miller | Pfizer, New York, NY, United States |

161 **Oncology Group**

| | |
|---|---|
| Tuomo Meretoja | Hospital District of Helsinki and Uusimaa, Helsinki, Finland |
| Heikki Joensuu | Hospital District of Helsinki and Uusimaa, Helsinki, Finland |
| Olli Carpén | Hospital District of Helsinki and Uusimaa, Helsinki, Finland |
| Johanna Mattson | Hospital District of Helsinki and Uusimaa, Helsinki, Finland |
| Eveliina Salminen | Hospital District of Helsinki and Uusimaa, Helsinki, Finland |
| Annika Auranen | Pirkanmaa Hospital District , Tampere, Finland |
| Peeter Karihtala | Northern Ostrobothnia Hospital District, Oulu, Finland |
| Päivi Auvinen | Northern Savo Hospital District, Kuopio, Finland |
| Klaus Elenius | Hospital District of Southwest Finland, Turku, Finland |
| Johanna Schleutker | Hospital District of Southwest Finland, Turku, Finland |
| Esa Pitkänen | Institute for Molecular Medicine Finland (FIMM), HiLIFE, University of Helsinki, Helsinki, Finland |
| Nina Mars | Institute for Molecular Medicine Finland (FIMM), HiLIFE, University of Helsinki, Helsinki, Finland |
| Mark Daly | Institute for Molecular Medicine Finland (FIMM), HiLIFE, University of Helsinki, Helsinki, Finland; Broad Institute of MIT and Harvard; Massachusetts General Hospital |
| Relja Popovic | Abbvie, Chicago, IL, United States |
| Jeffrey Waring | Abbvie, Chicago, IL, United States |
| Bridget Riley-Gillis | Abbvie, Chicago, IL, United States |
| Anne Lehtonen | Abbvie, Chicago, IL, United States |
| Jennifer Schutzman | Genentech, San Francisco, CA, United States |
| Julie Hunkapiller | Genentech, San Francisco, CA, United States |
| Natalie Bowers | Genentech, San Francisco, CA, United States |
| Rion Pendergrass | Genentech, San Francisco, CA, United States |
| Diptee Kulkarni | GlaxoSmithKline, Brentford, United Kingdom |
| Kirsi Auro | GlaxoSmithKline, Espoo, Finland |
| Alessandro Porello | Janssen Research & Development, LLC, Spring House, PA, United States |
| Andrey Loboda | Merck, Kenilworth, NJ, United States |
| Heli Lehtonen | Pfizer, New York, NY, United States |

| | |
|---|---|
| Stefan McDonough | Pfizer, New York, NY, United States |
| Sauli Vuoti | Janssen-Cilag Oy, Espoo, Finland |

**Opthalmology Group**



| | |
|---|---|
| Kai Kaarniranta | Northern Savo Hospital District, Kuopio, Finland |
| Joni A Turunen | Helsinki University Hospital and University of Helsinki, Helsinki, Finland; Eye Genetics Group, Folkhälsan Research Center, Helsinki, Finland |
| Terhi Ollila | Hospital District of Helsinki and Uusimaa, Helsinki, Finland |
| Hannu Uusitalo | Pirkanmaa Hospital District, Tampere, Finland |
| Juha Karjalainen | Institute for Molecular Medicine Finland (FIMM), HiLIFE, University of Helsinki, Helsinki, Finland |
| Esa Pitkänen | Institute for Molecular Medicine Finland (FIMM), HiLIFE, University of Helsinki, Helsinki, Finland |
| Mengzhen Liu | Abbvie, Chicago, IL, United States |
| Heiko Runz | Biogen, Cambridge, MA, United States |
| Stephanie Loomis | Biogen, Cambridge, MA, United States |
| Erich Strauss | Genentech, San Francisco, CA, United States |
| Natalie Bowers | Genentech, San Francisco, CA, United States |
| Hao Chen | Genentech, San Francisco, CA, United States |
| Rion Pendergrass | Genentech, San Francisco, CA, United States |

**Dermatology Group**

| | |
|---|---|
| Kaisa Tasanen | Northern Ostrobothnia Hospital District, Oulu, Finland |
| Laura Huilaja | Northern Ostrobothnia Hospital District, Oulu, Finland |
| Katariina Hannula-Jouppi | Hospital District of Helsinki and Uusimaa, Helsinki, Finland |
| Teea Salmi | Pirkanmaa Hospital District, Tampere, Finland |
| Sirkku Peltonen | Hospital District of Southwest Finland, Turku, Finland |
| Leena Koulu | Hospital District of Southwest Finland, Turku, Finland |
| Nizar Smaoui | Abbvie, Chicago, IL, United States |
| Fedik Rahimov | Abbvie, Chicago, IL, United States |
| Anne Lehtonen | Abbvie, Chicago, IL, United States |
| David Choy | Genentech, San Francisco, CA, United States |
| Rion Pendergrass | Genentech, San Francisco, CA, United States |

| | |
|---|---|
| Dawn Waterworth | Janssen Research & Development, LLC, Spring House, PA, United States |
| Kirsi Kalpala | Pfizer, New York, NY, United States |
| Ying Wu | Pfizer, New York, NY, United States |

**Odontology Group**

| | |
|---|---|
| Pirkko Pussinen | Hospital District of Helsinki and Uusimaa, Helsinki, Finland |
| Aino Salminen | Hospital District of Helsinki and Uusimaa, Helsinki, Finland |
| Tuula Salo | Hospital District of Helsinki and Uusimaa, Helsinki, Finland |
| David Rice | Hospital District of Helsinki and Uusimaa, Helsinki, Finland |
| Pekka Nieminen | Hospital District of Helsinki and Uusimaa, Helsinki, Finland |
| Ulla Palotie | Hospital District of Helsinki and Uusimaa, Helsinki, Finland |
| Maria Siponen | Northern Savo Hospital District, Kuopio, Finland |
| Liisa Suominen | Northern Savo Hospital District, Kuopio, Finland |
| Päivi Mäntylä | Northern Savo Hospital District, Kuopio, Finland |
| Ulvi Gursoy | Hospital District of Southwest Finland, Turku, Finland |
| Vuokko Anttonen | Northern Ostrobothnia Hospital District, Oulu, Finland |
| Kirsi Sipilä | Research Unit of Oral Health Sciences Faculty of Medicine, University of Oulu, Oulu, Finland; Medical Research Center, Oulu, Oulu University Hospital and University of Oulu, Oulu, Finland |
| Rion Pendergrass | Genentech, San Francisco, CA, United States |

163

**Women's Health and Reproduction Group**

| | |
|---|---|
| Hannele Laivuori | Institute for Molecular Medicine Finland (FIMM), HiLIFE, University of Helsinki, Helsinki, Finland |
| Venla Kurra | Pirkanmaa Hospital District, Tampere, Finland |
| Laura Kotaniemi-Talonen | Pirkanmaa Hospital District, Tampere, Finland |
| Oskari Heikinheimo | Hospital District of Helsinki and Uusimaa, Helsinki, Finland |
| Ilkka Kalliala | Hospital District of Helsinki and Uusimaa, Helsinki, Finland |
| Lauri Aaltonen | Hospital District of Helsinki and Uusimaa, Helsinki, Finland |
| Varpu Jokimaa | Hospital District of Southwest Finland, Turku, Finland |
| Johannes Kettunen | Northern Ostrobothnia Hospital District, Oulu, Finland |
| Marja Vääräsmäki | Northern Ostrobothnia Hospital District, Oulu, Finland |
| Outi Uimari | Northern Ostrobothnia Hospital District, Oulu, Finland |
| Laure Morin-Papunen | Northern Ostrobothnia Hospital District, Oulu, Finland |
| Maarit Niinimäki | Northern Ostrobothnia Hospital District, Oulu, Finland |

| | |
|---|---|
| Terhi Piltonen | Northern Ostrobothnia Hospital District, Oulu, Finland |
| Katja Kivinen | Institute for Molecular Medicine Finland (FIMM), HiLIFE, University of Helsinki, Helsinki, Finland |
| Elisabeth Widen | Institute for Molecular Medicine Finland (FIMM), HiLIFE, University of Helsinki, Helsinki, Finland |
| Taru Tukiainen | Institute for Molecular Medicine Finland (FIMM), HiLIFE, University of Helsinki, Helsinki, Finland |
| Mary Pat Reeve | Institute for Molecular Medicine Finland (FIMM), HiLIFE, University of Helsinki, Helsinki, Finland |
| Mark Daly | Institute for Molecular Medicine Finland (FIMM), HiLIFE, University of Helsinki, Helsinki, Finland; Broad Institute of MIT and Harvard; Massachusetts General Hospital |
| Niko Välimäki | University of Helsinki, Helsinki, Finland |
| Eija Laakkonen | University of Jyväskylä, Jyväskylä, Finland |
| Jaakko Tyrmi | University of Oulu, Oulu, Finland / University of Tampere, Tampere, Finland |
| Heidi Silven | University of Oulu, Oulu, Finland |
| Eeva Sliz | University of Oulu, Oulu, Finland |
| Riikka Arffman | University of Oulu, Oulu, Finland |
| Susanna Savukoski | University of Oulu, Oulu, Finland |
| Triin Laisk | Estonian biobank, Tartu, Estonia |
| Natalia Pujol | Estonian biobank, Tartu, Estonia |
| Mengzhen Liu | Abbvie, Chicago, IL, United States |
| Bridget Riley-Gillis | Abbvie, Chicago, IL, United States |
| Rion Pendergrass | Genentech, San Francisco, CA, United States |
| Janet Kumar | GlaxoSmithKline, Collegeville, PA, United States |
| Kirsi Auro | GlaxoSmithKline, Espoo, Finland |

**Depression Group**

| | |
|---|---|
| Iiris Hovatta | University of Helsinki, Finland |
| Chia-Yen Chen | Biogen, Cambridge, MA, United States |
| Erkki Isometsä | Hospital District of Helsinki and Uusimaa, Helsinki, Finland |
| Kumar Veerapen | Broad Institute, Cambridge, MA, United States |
| Hanna Ollila | Institute for Molecular Medicine Finland (FIMM), HiLIFE, University of Helsinki, Helsinki, Finland |
| Jaana Suvisaari | Finnish Institute for Health and Welfare (THL), Helsinki, Finland |
| Thomas Damm Als | Aarhus University, Denmark |

**ENT (ear, nose and throat) Group**

| | |
|---|---|
| Antti Mäkitie | Department of Otorhinolaryngology - Head and Neck Surgery, University of Helsinki and Helsinki University Hospital, Helsinki, Finland |
| Argyro Bizaki-Vallaskangas | Pirkanmaa Hospital District, Tampere, Finland |
| Sanna Toppila-Salmi | University of Helsinki, Finland |
| Tytti Willberg | Hospital District of Southwest Finland, Turku, Finland |
| Elmo Saarentaus | Institute for Molecular Medicine Finland (FIMM), HiLIFE, University of Helsinki, Helsinki, Finland |
| Antti Aarnisalo | Hospital District of Helsinki and Uusimaa, Helsinki, Finland |
| Eveliina Salminen | Hospital District of Helsinki and Uusimaa, Helsinki, Finland |
| Elisa Rahikkala | Northern Ostrobothnia Hospital District, Oulu, Finland |
| Johannes Kettunen | Northern Ostrobothnia Hospital District, Oulu, Finland |

**POI (premature ovarian failure) Group**

| | |
|---|---|
| Kristiina Aittomäki | Department of Medical Genetics, Helsinki University Central Hospital, Helsinki, Finland |

**LiverScore Group**

| | |
|---|---|
| Fredrik Åberg | Transplantation and Liver Surgery Clinic, Helsinki University Hospital, Helsinki University, Helsinki, Finland |

### FinnGen Analysis Working Group

| | |
|---|---|
| Mitja Kurki | Institute for Molecular Medicine Finland (FIMM), HiLIFE, University of Helsinki, Helsinki, Finland; Broad Institute, Cambridge, MA, United States |
| Samuli Ripatti | Institute for Molecular Medicine Finland (FIMM), HiLIFE, University of Helsinki, Helsinki, Finland |
| Mark Daly | Institute for Molecular Medicine, Finland (FIMM), HiLIFE, University of Helsinki, Helsinki, Finland; Broad Institute of MIT and Harvard; Massachusetts General Hospital |
| Juha Karjalainen | Institute for Molecular Medicine Finland (FIMM), HiLIFE, University of Helsinki, Helsinki, Finland |
| Aki Havulinna | Institute for Molecular Medicine Finland (FIMM), HiLIFE, University of Helsinki, Helsinki, Finland; Finnish Institute for Health and Welfare (THL), Helsinki, Finland |
| Juha Mehtonen | Institute for Molecular Medicine Finland (FIMM), HiLIFE, University of Helsinki, Helsinki, Finland |
| Priit Palta | Institute for Molecular Medicine Finland (FIMM), HiLIFE, University of Helsinki, Helsinki, Finland |
| Shabbeer Hassan | Institute for Molecular Medicine Finland (FIMM), HiLIFE, University of Helsinki, Helsinki, Finland |

| | |
|---|---|
| Pietro Della Briotta Parolo | Institute for Molecular Medicine Finland (FIMM), HiLIFE, University of Helsinki, Helsinki, Finland |
| Wei Zhou | Broad Institute, Cambridge, MA, United States |
| Mutaamba Maasha | Broad Institute, Cambridge, MA, United States |
| Kumar Veerapen | Broad Institute, Cambridge, MA, United States |
| Shabbeer Hassan | Institute for Molecular Medicine Finland (FIMM), HiLIFE, University of Helsinki, Helsinki, Finland |
| Susanna Lemmelä | Institute for Molecular Medicine Finland (FIMM), HiLIFE, University of Helsinki, Helsinki, Finland |
| Manuel Rivas | University of Stanford, Stanford, CA, United States |
| Mari E. Niemi | Institute for Molecular Medicine Finland (FIMM), HiLIFE, University of Helsinki, Helsinki, Finland |
| Aarno Palotie | Institute for Molecular Medicine Finland (FIMM), HiLIFE, University of Helsinki, Helsinki, Finland |
| Aoxing Liu | Institute for Molecular Medicine Finland (FIMM), HiLIFE, University of Helsinki, Helsinki, Finland |
| Arto Lehisto | Institute for Molecular Medicine Finland (FIMM), HiLIFE, University of Helsinki, Helsinki, Finland |
| Andrea Ganna | Institute for Molecular Medicine Finland (FIMM), HiLIFE, University of Helsinki, Helsinki, Finland |
| Vincent Llorens | Institute for Molecular Medicine Finland (FIMM), HiLIFE, University of Helsinki, Helsinki, Finland |
| Hannele Laivuori | Institute for Molecular Medicine Finland (FIMM), HiLIFE, University of Helsinki, Helsinki, Finland |
| Taru Tukiainen | Institute for Molecular Medicine Finland (FIMM), HiLIFE, University of Helsinki, Helsinki, Finland |
| Mary Pat Reeve | Institute for Molecular Medicine Finland (FIMM), HiLIFE, University of Helsinki, Helsinki, Finland |
| Henrike Heyne | Institute for Molecular Medicine Finland (FIMM), HiLIFE, University of Helsinki, Helsinki, Finland |
| Nina Mars | Institute for Molecular Medicine Finland (FIMM), HiLIFE, University of Helsinki, Helsinki, Finland |
| Joel Rämö | Institute for Molecular Medicine Finland (FIMM), HiLIFE, University of Helsinki, Helsinki, Finland |
| Elmo Saarentaus | Institute for Molecular Medicine Finland (FIMM), HiLIFE, University of Helsinki, Helsinki, Finland |
| Hanna Ollila | Institute for Molecular Medicine Finland (FIMM), HiLIFE, University of Helsinki, Helsinki, Finland |
| Rodos Rodosthenous | Institute for Molecular Medicine Finland (FIMM), HiLIFE, University of Helsinki, Helsinki, Finland |
| Satu Strausz | Institute for Molecular Medicine Finland (FIMM), HiLIFE, University of Helsinki, Helsinki, Finland |
| Tuula Palotie | University of Helsinki and Hospital District of Helsinki and Uusimaa, Helsinki, Finland |
| Kimmo Palin | University of Helsinki, Helsinki, Finland |
| Javier Garcia-Tabuenca | University of Tampere, Tampere, Finland |
| Harri Siirtola | University of Tampere, Tampere, Finland |
| Tuomo Kiiskinen | Institute for Molecular Medicine Finland (FIMM), HiLIFE, University of Helsinki, Helsinki, Finland |
| Jiwoo Lee | Institute for Molecular Medicine Finland (FIMM), HiLIFE, University of Helsinki, Helsinki, Finland; Broad Institute, Cambridge, MA, United States |
| Kristin Tsuo | Institute for Molecular Medicine Finland (FIMM), HiLIFE, University of Helsinki, Helsinki, Finland; Broad Institute, Cambridge, MA, United States |

| | |
|---|---|
| Amanda Elliott | Institute for Molecular Medicine Finland (FIMM), HiLIFE, University of Helsinki, Helsinki, Finland; Broad Institute, Cambridge, MA, USA and Massachusetts General Hospital, Boston, MA, USA |
| Kati Kristiansson | THL Biobank / Finnish Institute for Health and Welfare (THL), Helsinki, Finland |
| Mikko Arvas | Finnish Red Cross Blood Service / Finnish Hematology Registry and Clinical Biobank, Helsinki, Finland |
| Kati Hyvärinen | Finnish Red Cross Blood Service, Helsinki, Finland |
| Jarmo Ritari | Finnish Red Cross Blood Service, Helsinki, Finland |
| Olli Carpén | Helsinki Biobank / Helsinki University and Hospital District of Helsinki and Uusimaa, Helsinki |
| Johannes Kettunen | Northern Finland Biobank Borealis / University of Oulu / Northern Ostrobothnia Hospital District, Oulu, Finland |
| Katri Pylkäs | University of Oulu, Oulu, Finland |
| Eeva Sliz | University of Oulu, Oulu, Finland |
| Minna Karjalainen | University of Oulu, Oulu, Finland |
| Tuomo Mantere | Northern Finland Biobank Borealis / University of Oulu / Northern Ostrobothnia Hospital District, Oulu, Finland |
| Eeva Kangasniemi | Finnish Clinical Biobank Tampere / University of Tampere / Pirkanmaa Hospital District, Tampere, Finland |
| Sami Heikkinen | University of Eastern Finland, Kuopio, Finland |
| Arto Mannermaa | Biobank of Eastern Finland / University of Eastern Finland / Northern Savo Hospital District, Kuopio, Finland |
| Eija Laakkonen | University of Jyväskylä, Jyväskylä, Finland |
| Nina Pitkänen | Auria Biobank / University of Turku / Hospital District of Southwest Finland, Turku, Finland |
| Samuel Lessard | Translational Sciences, Sanofi R&D, Framingham, MA, USA |
| Clément Chatelain | Translational Sciences, Sanofi R&D, Framingham, MA, USA |


### Biobank directors


| | |
|---|---|
| Perttu Terho | Auria Biobank / University of Turku / Hospital District of Southwest Finland, Turku, Finland |
| Sirpa Soini | THL Biobank / Finnish Institute for Health and Welfare (THL), Helsinki, Finland |
| Jukka Partanen | Finnish Red Cross Blood Service / Finnish Hematology Registry and Clinical Biobank, Helsinki, Finland |
| Eero Punkka | Helsinki Biobank / Helsinki University and Hospital District of Helsinki and Uusimaa, Helsinki |
| Raisa Serpi | Northern Finland Biobank Borealis / University of Oulu / Northern Ostrobothnia Hospital District, Oulu, Finland |
| Sanna Siltanen | Finnish Clinical Biobank Tampere / University of Tampere / Pirkanmaa Hospital District, Tampere, Finland |
| Veli-Matti Kosma | Biobank of Eastern Finland / University of Eastern Finland / Northern Savo Hospital District, Kuopio, Finland |

| | |
|---|---|
| Teijo Kuopio | Central Finland Biobank / University of Jyväskylä / Central Finland Health Care District, Jyväskylä, Finland |


### FinnGen Teams

**Administration**

| | |
|---|---|
| Anu Jalanko | Institute for Molecular Medicine Finland (FIMM), HiLIFE, University of Helsinki, Helsinki, Finland |
| Huei-Yi Shen | Institute for Molecular Medicine Finland (FIMM), HiLIFE, University of Helsinki, Helsinki, Finland |
| Risto Kajanne | Institute for Molecular Medicine Finland (FIMM), HiLIFE, University of Helsinki, Helsinki, Finland |
| Mervi Aavikko | Institute for Molecular Medicine Finland (FIMM), HiLIFE, University of Helsinki, Helsinki, Finland |

**Analysis**

| | |
|---|---|
| Mitja Kurki | Institute for Molecular Medicine Finland (FIMM), HiLIFE, University of Helsinki, Helsinki, Finland; Broad Institute, Cambridge, MA, United States |
| Juha Karjalainen | Institute for Molecular Medicine Finland (FIMM), HiLIFE, University of Helsinki, Helsinki, Finland |
| Pietro Della Briotta Parolo | Institute for Molecular Medicine Finland (FIMM), HiLIFE, University of Helsinki, Helsinki, Finland |
| Arto Lehisto | Institute for Molecular Medicine Finland (FIMM), HiLIFE, University of Helsinki, Helsinki, Finland |
| Juha Mehtonen | Institute for Molecular Medicine Finland (FIMM), HiLIFE, University of Helsinki, Helsinki, Finland |
| Wei Zhou | Broad Institute, Cambridge, MA, United States |
| Masahiro Kanai | Broad Institute, Cambridge, MA, United States |
| Mutaamba Maasha | Broad Institute, Cambridge, MA, United States |
| Kumar Veerapen | Broad Institute, Cambridge, MA, United States |

**Clinical Endpoint Development**

| | |
|---|---|
| Hannele Laivuori | Institute for Molecular Medicine Finland (FIMM), HiLIFE, University of Helsinki, Helsinki, Finland |
| Aki Havulinna | Institute for Molecular Medicine Finland (FIMM), HiLIFE, University of Helsinki, Helsinki, Finland; Finnish Institute for Health and Welfare (THL), Helsinki, Finland |
| Susanna Lemmelä | Institute for Molecular Medicine Finland (FIMM), HiLIFE, University of Helsinki, Helsinki, Finland |
| Tuomo Kiiskinen | Institute for Molecular Medicine Finland (FIMM), HiLIFE, University of Helsinki, Helsinki, Finland |

L. Elisa Lahtela Institute for Molecular Medicine Finland (FIMM), HiLIFE, University of Helsinki, Helsinki, Finland

**Communication**

Mari Kaunisto Institute for Molecular Medicine Finland (FIMM), HiLIFE, University of Helsinki, Helsinki, Finland

**E-Science**

Elina Kilpeläinen Institute for Molecular Medicine Finland (FIMM), HiLIFE, University of Helsinki, Helsinki, Finland

Timo P. Sipilä Institute for Molecular Medicine Finland (FIMM), HiLIFE, University of Helsinki, Helsinki, Finland

Oluwaseun Alexander Dada Institute for Molecular Medicine Finland (FIMM), HiLIFE, University of Helsinki, Helsinki, Finland

Awaisa Ghazal Institute for Molecular Medicine Finland (FIMM), HiLIFE, University of Helsinki, Helsinki, Finland

Anastasia Kytölä Institute for Molecular Medicine Finland (FIMM), HiLIFE, University of Helsinki, Helsinki, Finland

Rigbe Weldatsadik Institute for Molecular Medicine Finland (FIMM), HiLIFE, University of Helsinki, Helsinki, Finland

**Genotyping**

Kati Donner Institute for Molecular Medicine Finland (FIMM), HiLIFE, University of Helsinki, Helsinki, Finland

Timo P. Sipilä Institute for Molecular Medicine Finland (FIMM), HiLIFE, University of Helsinki, Helsinki, Finland

**Sample Collection Coordination**

Anu Loukola Helsinki Biobank / Helsinki University and Hospital District of Helsinki and Uusimaa, Helsinki

**Sample Logistics**

Päivi Laiho THL Biobank / Finnish Institute for Health and Welfare (THL), Helsinki, Finland

Tuuli Sistonen THL Biobank / Finnish Institute for Health and Welfare (THL), Helsinki, Finland

Essi Kaiharju THL Biobank / Finnish Institute for Health and Welfare (THL), Helsinki, Finland

Markku Laukkanen THL Biobank / Finnish Institute for Health and Welfare (THL), Helsinki, Finland

Elina Järvensivu THL Biobank / Finnish Institute for Health and Welfare (THL), Helsinki, Finland

Sini Lähteenmäki THL Biobank / Finnish Institute for Health and Welfare (THL), Helsinki, Finland

Lotta Männikkö THL Biobank / Finnish Institute for Health and Welfare (THL), Helsinki, Finland

Regis Wong THL Biobank / Finnish Institute for Health and Welfare (THL), Helsinki, Finland

Auli Toivola THL Biobank / Finnish Institute for Health and Welfare (THL), Helsinki, Finland

**Registry Data Operations**

| | |
|---|---|
| Minna Brunfeldt | THL Biobank / Finnish Institute for Health and Welfare (THL), Helsinki, Finland |
| Hannele Mattsson | THL Biobank / Finnish Institute for Health and Welfare (THL), Helsinki, Finland |
| Kati Kristiansson | THL Biobank / Finnish Institute for Health and Welfare (THL), Helsinki, Finland |
| Susanna Lemmelä | Institute for Molecular Medicine Finland (FIMM), HiLIFE, University of Helsinki, Helsinki, Finland |
| Sami Koskelainen | THL Biobank / Finnish Institute for Health and Welfare (THL), Helsinki, Finland |
| Tero Hiekkalinna | THL Biobank / Finnish Institute for Health and Welfare (THL), Helsinki, Finland |
| Teemu Paajanen | THL Biobank / Finnish Institute for Health and Welfare (THL), Helsinki, Finland |

**Sequencing Informatics**

| | |
|---|---|
| Priit Palta | Institute for Molecular Medicine Finland (FIMM), HiLIFE, University of Helsinki, Helsinki, Finland |
| Kalle Pärn | Institute for Molecular Medicine Finland (FIMM), HiLIFE, University of Helsinki, Helsinki, Finland |
| Mart Kals | Institute for Molecular Medicine Finland (FIMM), HiLIFE, University of Helsinki, Helsinki, Finland |
| Shuang Luo | Institute for Molecular Medicine Finland (FIMM), HiLIFE, University of Helsinki, Helsinki, Finland |
| Vishal Sinha | Institute for Molecular Medicine Finland (FIMM), HiLIFE, University of Helsinki, Helsinki, Finland |

**Trajectory**

| | |
|---|---|
| Tarja Laitinen | Pirkanmaa Hospital District, Tampere, Finland |
| Mary Pat Reeve | Institute for Molecular Medicine Finland (FIMM), HiLIFE, University of Helsinki, Helsinki, Finland |
| Marianna Niemi | University of Tampere, Tampere, Finland |
| Kumar Veerapen | Broad Institute, Cambridge, MA, United States |
| Harri Siirtola | University of Tampere, Tampere, Finland |
| Javier Gracia-Tabuenca | University of Tampere, Tampere, Finland |
| Mika Helminen | University of Tampere, Tampere, Finland |
| Tiina Luukkaala | University of Tampere, Tampere, Finland |
| Iida Vähätalo | University of Tampere, Tampere, Finland |

**Data protection officer**

| | |
|---|---|
| Jyrki Pitkänen | Institute for Molecular Medicine Finland (FIMM), HiLIFE, University of Helsinki, Helsinki, Finland |

**FINBB - Finnish biobank cooperative**

| | |
|---|---|
| Marco Hautalahti | Finnish Biobank Cooperative - FINBB |
| Johanna Mäkelä | Finnish Biobank Cooperative - FINBB |
| Sarah Smith | Finnish Biobank Cooperative - FINBB |
| Tom Southerington | Finnish Biobank Cooperative - FINBB |
